# Comparative effectiveness of the BNT162b2 vs ChAdOx1 vaccine against Covid-19

**DOI:** 10.1101/2021.12.18.21268039

**Authors:** Junqing xie, Shuo Feng, Xintong Li, Ester Gea-Mallorquí, Albert Prats-Uribe, Dani Prieto-Alhambra

## Abstract

Although pivotal trials with varying populations and study methods suggest higher efficacy for mRNA than adenoviral Covid-19 vaccines, no direct evidence is available. Here, we conducted a head-to-head comparison of BNT162b2 versus ChAdOx1 against Covid-19. We analysed 235,181 UK Biobank participants aged 50 years or older and vaccinated with one or two doses of BNT162b2 or ChAdOx1. People were followed from the vaccination date until 18/10/2021. Inverse probability weighting was used to minimise confounding and the Cox models to derive hazard ratio. We found that, compared with two doses of ChAdOx1, vaccination with BNT162b2 was associated with 30% lower risks of both SARS-CoV-2 infection and related hospitalisation during the period dominated by the delta variant. Also, this comparative effectiveness was consistent across several subgroups and persisted for at least six months, suggesting no differential waning between the two vaccines. Our findings can inform evidence-based Covid-19 vaccination campaigns and booster strategies.

## Introduction

To date, four vaccines against the Coronavirus disease 2019 (Covid-19) have been approved for use in the UK by the Medicines and Healthcare products Regulatory Agency: the Pfizer-BioNTech BNT162b2, Moderna’s mRNA-1273, Oxford-AstraZeneca’s ChAdOx1, and Janssen’s Ad26.CoV2.S. Although phase 3 trials suggested that all four have high clinical efficacy, mRNA vaccines demonstrated numerically greater efficacy than adenoviral-based ones: BNT162b2 reported 95% efficacy against symptomatic covid-19,^1^ mRNA-1273 94.1% efficacy,^2^ whilst ChAdOx1 had 70.4%,^3^ and Ad26.CoV2.S had 66.9% efficacy.^4^ However, notable differences in study designs made it difficult to compare vaccine efficacy based on these trials, including different populations recruited in different regions and at different times, diverse primary endpoint definitions, and heterogeneous statistical analysis methods.

Specific for the two more widely utilized vaccines, BNT162b2 and ChAdOx1, several observational studies have recently evaluated their effectiveness in multiple real-world settings from Israel,^5^ the UK,^6^ and Spain,^7^ amongst others. Although useful, none of these studies conducted head-to-head comparisons of vaccine effectiveness. With the ongoing pandemic and rapid rollout of the Covid-19 vaccination programme all over the world, evidence on their comparative performance has become more crucial to inform policy decisions on optimizing vaccine implementation strategies, not only in countries where greater coverage of the prime dose is urgently required or in countries where boost doses and sequential immunization are being considered. Therefore, this study aimed to estimate the comparative effectiveness of BNT162b2 vs ChAdOx1 vaccines against Covid-19 infection and hospitalization in a large and rich prospective cohort of people aged 50 years or older.

## Results

### Data linkage and study cohorts

During the one-dose enrolment period, 70,097 and 98,551 people received the first BNT162b2 and ChAdOx1 Covid-19 vaccines, respectively. These figures were 67,813 and 89,030 accordingly for people receiving the second dose during the two-dose enrolment period. Vaccine uptake over calendar time in our study population is depicted in **Figure 1**.

**Figure 1:**
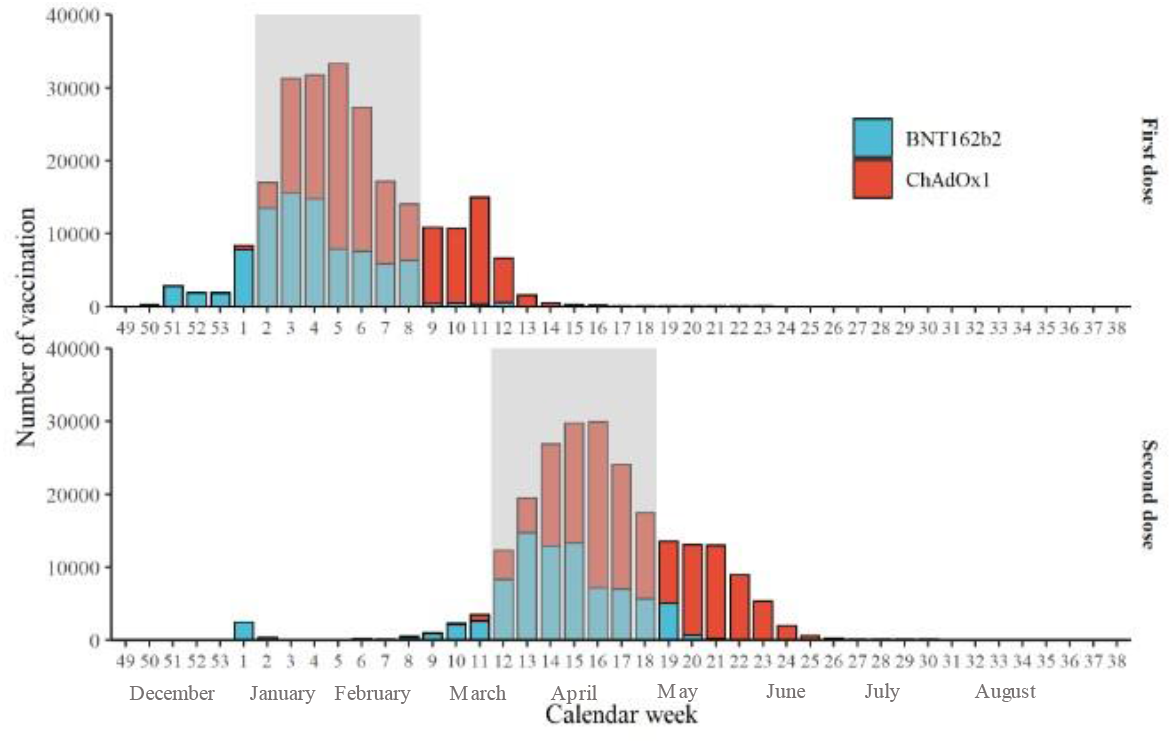
The number of people receiving either BNT162b2 or ChAdOx1 Covid-19 vaccine in the study cohort from Dec 01, 2020 to Sep 21, 2021. The rectangle background with grey color depicts the vaccination anchor windows. Only people who received study vaccines within the anchor windows were included for the comparative analysis. We defined the 2^rd^ to 8^th^ (Jan 11, 2021 to Feb 28, 2021) and 12^th^ to 18^th^ (March 22, 2021 to May 9, 2021) calendar weeks as the two anchor windows for the one-dose and two-dose cohorts, respectively. The decision-making for these anchor windows was based on (1) there were both BNT162b2 and ChAdOx1 vaccines delivered in each epidemiological week of the window, (2) numbers of the two vaccines were generally comparable, and (3) UK’s policy on the gap between the first and second dose was 10 to 12 weeks for both vaccines.

### Baseline characteristics

In the one-dose vaccine cohorts, people receiving BNT162b2 were slightly older (mean (sd) age: 71.35 (7.21) years) than those receiving ChAdOx1 (mean (sd) age: 71.06 (6.02) years). Sex (44.5% vs 44.1% male) and ethnicity (91.2% vs 92.6% White) were comparable between the two groups. Little difference was seen in the prevalence of medicines or comorbidities. The main differences between cohorts were vaccination dates and socio-economic factors such as income (**Figure 2** and **Supplementary Table 2**). Similar patterns of baseline characteristic differences were also seen in the two-dose comparison cohorts (**Figure 2 and Supplementary Table 3**). All covariates were balanced after IPW weighting with an absolute standardised mean difference < 0.1, including the date of vaccination.

**Figure 2:**
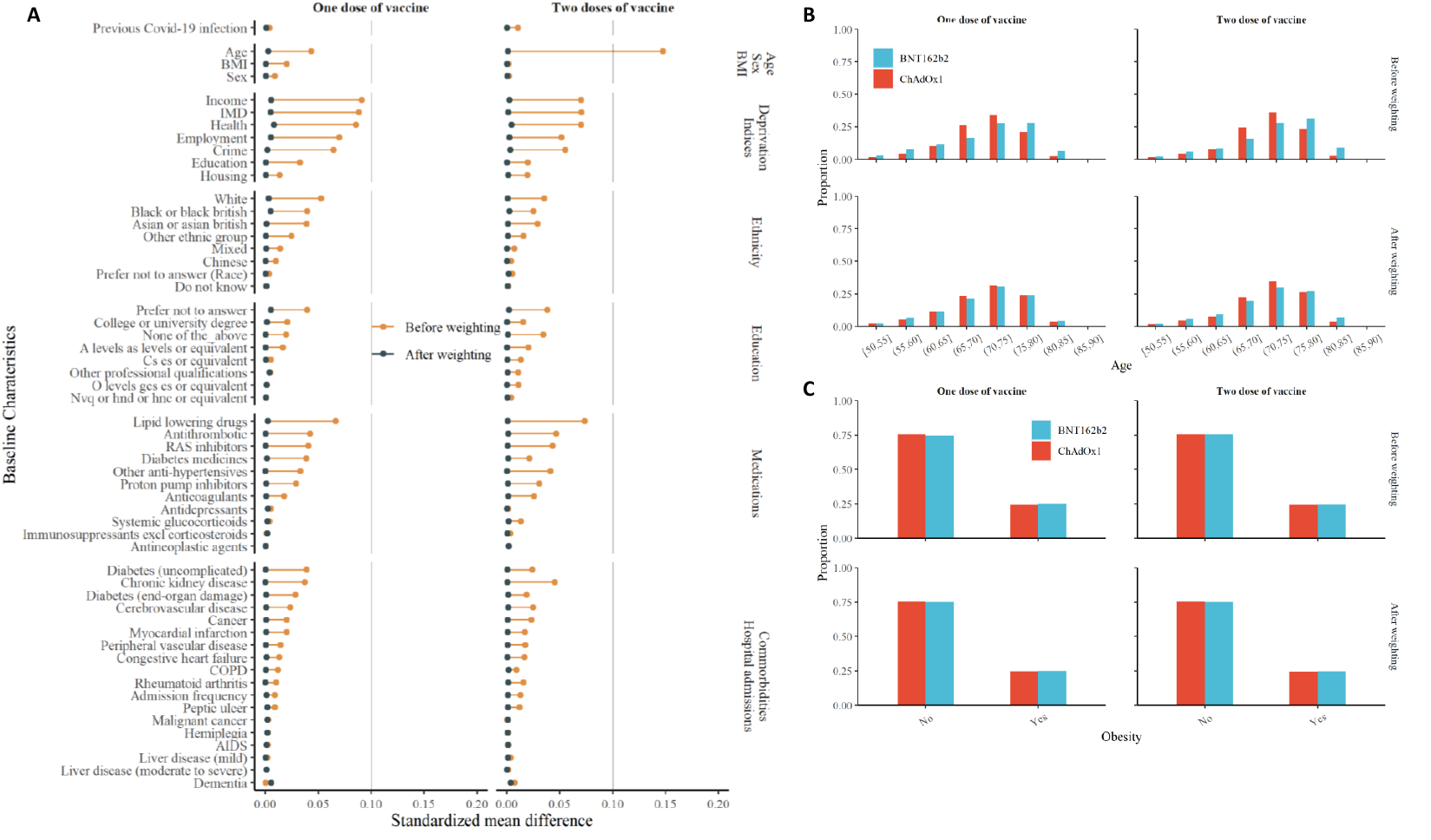
Balance of baseline covariates for the one dose and two doses cohorts, before and after weighting. Notes: (A) standardized mean difference for all covariates included in the propensity score. (B, C) distribution of age and % obesity according to vaccine received

### Incidence and hazard ratio

Over the 14,630 and 20,714 person-years of follow-up for the one dose BNT162b2 and ChAdOx1 recipient, 200 and 261 people tested positive for SARS-CoV-2, equivalent to incidence rates (IR) of 13.7 and 12.6/1,000 person-years, respectively, and an unadjusted hazard ratio (HR) of 1.08 (95% 0.90 - 1.30). After inverse probability weighting, the HR changed to 0.72 (95% 0.58 - 0.88), favouring BNT162b2 in the overall population (**Table 1**). In contrast, the incidences of Covid-19 hospitalisation were similar among the one dose BNT162b2 (IR: 3.07 per 1,000 person-years) and ChAdOx1 (IR: 2.17 per 1,000 person-years) cohorts, with no noticeable differences between both vaccine groups: adjusted/weighted HR 0.87 (95% 0.53 - 1.41).

**Table 1:** Incidence rate and hazard ratio of COIVD-19 infection and hospitalisation following one and two doses of vaccines.

|  | BNT162b2 |  |  | ChAdOx1 |  |  |  |  |
| --- | --- | --- | --- | --- | --- | --- | --- | --- |
|  | Vaccination<br>years | Cases | Rate, per<br>1000 PYs | Vaccination<br>years | Cases | Rate, per<br>1000 PYs | Unadjusted<br>hazard ratio | Adjusted hazard<br>ratio |
| One dose |  |  |  |  |  |  |  |  |
| Covid-19 infection | 14630 | 200 | 13.7 | 20714 | 261 | 12.6 | 1.08 (0.90 - 1.30) | 0.72 (0.58 - 0.88) |
| Covid-19 hospitalisation | 14654 | 45 | 3.07 | 20746 | 45 | 2.17 | 1.41 (0.95 - 2.09) | 0.87 (0.53 - 1.41) |
| Two doses |  |  |  |  |  |  |  |  |
| Covid-19 infection | 34991 | 1361 | 38.9 | 44084 | 2497 | 56.6 | 0.72 (0.57 - 0.91) | 0.70 (0.65 - 0.75) |
| Covid-19 hospitalisation | 35176 | 122 | 3.47 | 44421 | 202 | 4.55 | 0.66 (0.62 - 0.71) | 0.71 (0.55 - 0.90) |

PYs: person-years

After the second dose, 1,361/34,991 person-years (IR: 38.9 per 1,000 person-years) and 2,497/44,084 person-years (IR: 56.6 per 1,000 person-years) were identified positive for SARS-CoV-2 among BNT162b2 and ChAdOx1 recipients respectively. Unadjusted (0.72, 95% 0.57 - 0.91) and adjusted HR (0.70, 95% 0.65 - 0.75) were almost identical, favouring BNT162b2 (**Table 1**). The rates of Covid-19 hospitalisation remained low in both cohorts, but higher amongst ChAdOx1 (IR: 4.55 per 1,000 person-years) compared to BNT162b2 recipients (IR: 3.47 per 1,000 person-years), with adjusted HR of 0.71 (95% 0.55 - 0.90) favouring BNT162b2.

### Kaplan–Meier curve of Covid-19 infection and hospitalisation

Kaplan–Meier curves stratified by vaccine depicted similar trends between Covid-19 infection and hospitalisation. The cumulative incidence after the first dose increased rapidly in the early follow-up but flattened later until 14 weeks after vaccination (**Figure 3**), corresponding to the calendar period from January to March 2021 (**Supplementary Figure 1**). Conversely, the trend of cumulative incidence was reversed for the two-dose cohorts, with a substantial increase starting 12 weeks after the second dose (Figure 3), corresponding to the calendar period from June to October 2021 (**Supplementary Figure 1**). Changes in community transmission over time in the general population of England and among UK Biobank participants are shown in **Supplementary Figure 1**.

**Figure 3:**
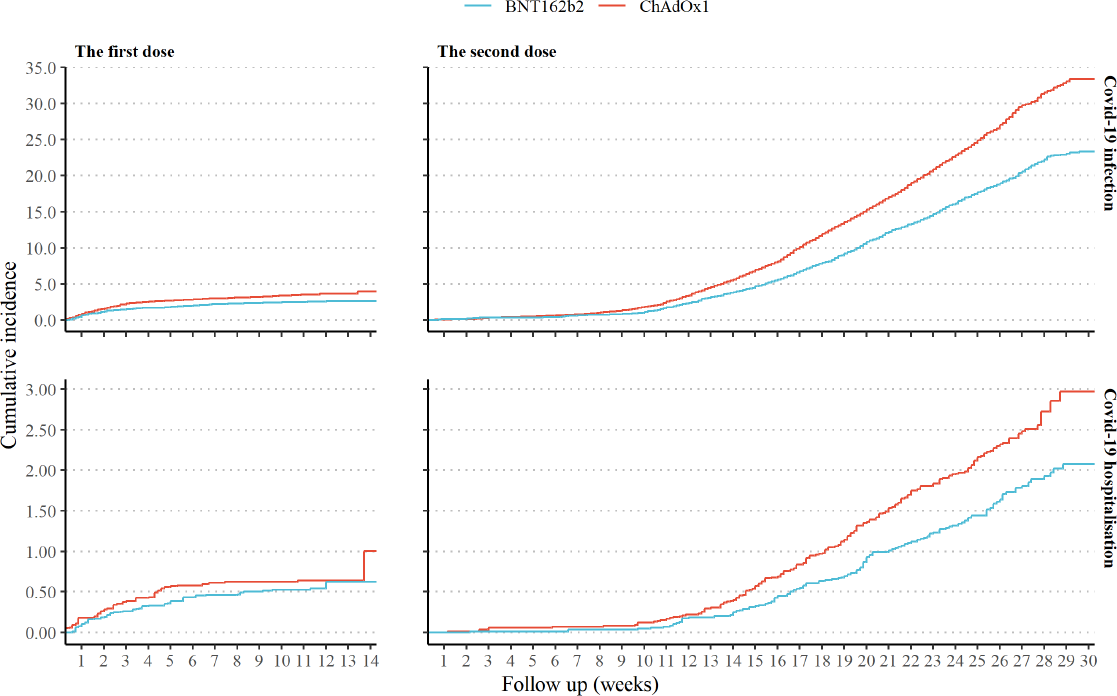
Kaplan-Meier cumulative incidence curves of Covid-19 infection and hospitalization after one and two doses of vaccines. Notes: The different Y-axis scales for Covid-19 infection and hospitalisation outcomes. The longest follow-up was 14 weeks after the first dose and 30 weeks after the second dose of the vaccine.

### Comparative effectiveness across sub-populations and over-time

The risk of Covid-19 infection was consistently lower in people receiving two doses of BNT162b2 compared to those receiving two doses of ChAdOx1 across all strata. HRs ranged from 0.65 (95% 0.59 - 0.71) for females to 0.80 (95% 0.70 - 0.90) for oldest-old adults.

Notably, incidence rates of Covid-19 infection increased substantially during the study period, from 3.93 and 4.87 per 1,000 person-years at the 0–4-week window to 74.74 and 111.87 per 1,000 person-years at the >24-week window in BNT162b2 and ChAdOx1 cohorts, respectively. However, as reflected by HRs, the comparative risks were stable for at least six months (**Figure 4**). Although a similar pattern was observed for Covid-19 hospitalisation, power was limited for the time-split analysis, in particular for the 0–4-week window (HR: 0.16, 95% 0.02 - 1.54) and 4–8-week window (HR: 2.48, 95% 0.16 - 39.66), due to the extremely rare events observed in the first few weeks after the second vaccination (**Figure 4**).

**Figure 4:**
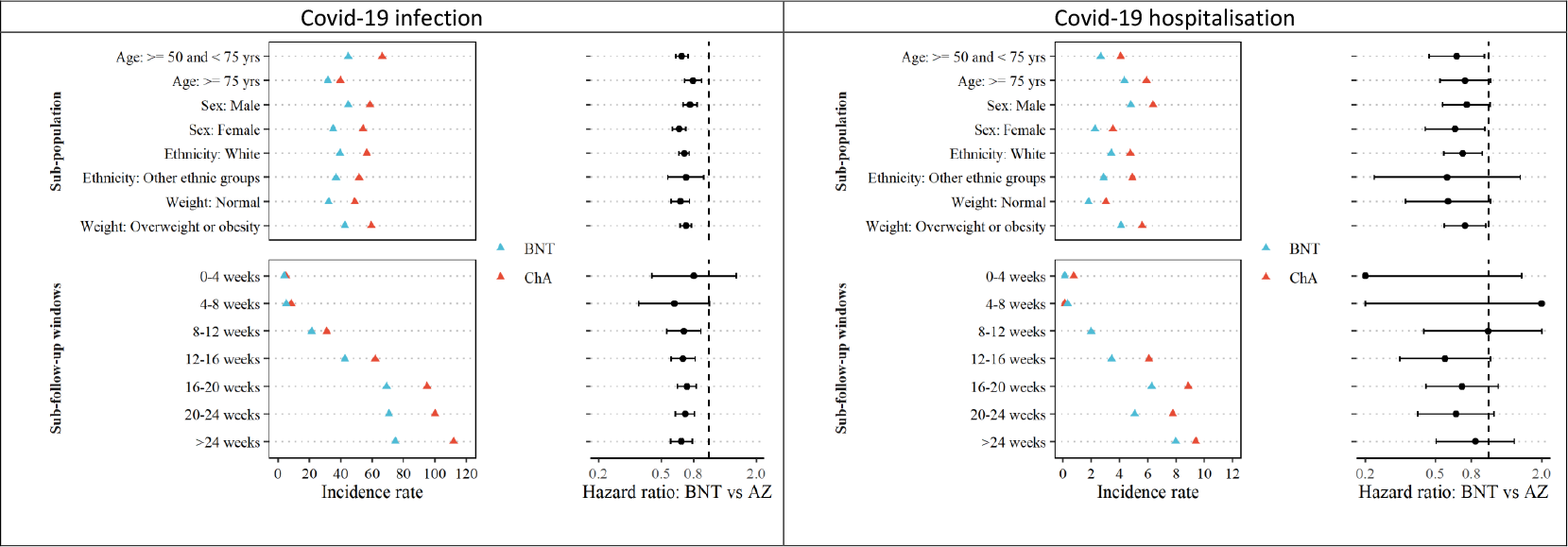
Comparative effectiveness of two doses BNT162b2 vs ChAdOx1 vaccine on Covid-19 infection and hospitalisation across key subgroups and over time. Notes: The different X-axis range of incidence rate for the Covid-19 infection and hospitalisation outcomes (unit, per 1000 person-years). Hazard ratios for Covid-19 hospitalisation were truncated at 0.2 (lowest) and 2 (highest).

### Negative control and sensitivity analyses

Adjusted hazard ratios for proposed negative clinical outcomes according to vaccine received among the one dose cohort were 1.01 (0.95 to 1.17) for limb pain, 0.93 (0.82 to 1.05) for fracture, and 0.88 (0.51 to 1.51) for peptic ulcer.

The main results were consistent in the propensity score-matched cohorts with slightly wider confidence intervals (**Supplementary Table 4**).

## Discussion

Among adults aged 50 years and older, we found that people receiving two doses of BNT162b2 vaccine had 30% lower risks of both Covid-19 diagnosis and related hospital admission, compared with those vaccinated with ChAdOx1. A similar difference was also observed among the one dose recipients, yet, only for Covid-19 diagnosis. Notably, the two-dose comparative effectiveness was consistent across several high-risk groups, such as oldest-old,^7^ male,^8^ ethnic minority^9^ and those with overweight or obesity,^10^ and persisted over time for over six months.

To date, only two Phase 2 randomized controlled trial has directly compared the efficacy of mRNA with adenovirus-based Covid-19 vaccines by using immunogenic endpoints. Liu et al.^11^ found that the BNT162B2-BNT162B2 appeared more immunogenic than the ChAdOx1-ChAdOx1 schedule, with higher levels of antibodies at 28 days after the first and second dose. A similar finding was reported in the Borobia et al. study comparing the heterologous BNT162B2-ChAdOx1 and the homogenous ChAdOx1-ChAdOx1 vaccine regimens.^12,13^ In line with this evidence, our study showed a lower risk of Covid-19 outcomes in people vaccinated with BNT162b2 overall and across subgroups.

However, a recent preprint study by Hulme et al.^14^ showed no difference among recipients of these two vaccines regarding the risk of SARS-CoV-2 positive test, COVID-19 related accident & emergency attendance and hospital admission. Several fundamental factors might explain the disagreement with our results. First, Hulme’s study only included health and social care workers who are more likely to be exposed to SARS-CoV-2 than our cohort: a community-based “healthy volunteer” population,^15^ which was corroborated by the higher infection rate in their study (58 per 1,000 person-years) compared to ours (∼13 per 1,000 person-years) during the first few weeks following the first dose. Second, both our and Hulme’s studies found that, on average, vaccination with BNT162B2 happened earlier than ChAdOx1. Given the variation over time in community transmission in England (**Supplementary Figure 1**), insufficient control for vaccination date can result in an inflated risk of COVID-19 amongst BNT162B2 recipients. Informed by this, we observed a substantial change of hazard ratio after the complete alignment of the first vaccination date by weighting in our study. Thirdly, Hulme’s risk assessment period started from the first dose (January to February 2021) and ended on 13 June 2021, reflecting a mixed effect of one and two doses of vaccines.

Finally, our data for the first time demonstrated that the observed differences did not attenuate over time, verifying the hypothesis from a few recent preprint studies.^16,17^

### Strengths and limitations

The leading challenges in estimating vaccine effectiveness with observational data lies in confounding by indication and potentially differential testing rates between exposed vaccinated and unvaccinated populations.^18,19^ However, our study minimized the impact of such differences by comparing two vaccines and restricting the analysis to a period when both vaccines were available and had a similar national delivery. UK data are ideal for comparative effectiveness research into Covid-19 vaccines, as both BNT162b2 and ChAdOx1 were rolled out simultaneously for the target population (adults &#x2265; 50 years) included in our analyses.^20 21,22^.

However, additional limitations remain. Information on a few participants’ characteristics was collected ten years ago and may have changed since then. However, given that all people at the cohort recruitment were already middle-aged or older adults (40-69 years old), we expected any changes in those features such as socio-economic status and education are likely to be minor or unrelated to the choice of vaccine types. Admittedly, misclassification of covariates could bias the genuine association towards the null and lead to underestimating the comparative vaccine effectiveness. Our study has several unique strengths. First, the granularity of UK Biobank data and comprehensive linkage to external data sources allowed us to measure and control for an extensive array of confounders, including demographics, socio-economic deprivation, comorbidity, and medication usage. Our negative control outcome analyses provided reassurance that no significant residual confounding remained after adjusting the mentioned covariates using IPW methods.

Secondly, the proposed study outcomes (Covid-19 and related hospitalization) were identified through linkage to official national databases of tests and hospital inpatient data, minimising the risk of misclassification. Finally, the sample size of our cohort triplicated that of the largest phase III trials, enabling us to detect differences in hospital admission rates, which seemed underpowered in randomized trials.

### Conclusions

Our findings support evidence from pivotal trials suggesting that BNT162b2 provides additional protection against Covid-19 and hospitalisation than ChAdOx1 vaccination. For the first time, we demonstrated that this comparative effectiveness endured over six months when the Delta variant was predominant, and community transmission kept increasing in the UK. These findings highlight the importance of continuous monitoring of the effectiveness of different vaccines against emerging SARS-CoV-2 variants to inform future booster campaigns and vaccine combinations strategies.

## Methods

### Study population and design

We used data from the UK Biobank (UKBB) cohort, a prospective study of approximately 500,000 individuals aged between 40 and 69 at baseline recruited in 2007-2010 from England (89%), Scotland (7%) and Wales (4%). All participants provided comprehensive information on socio-demographic, lifestyle, and health-related factors, which has been described in detail elsewhere.^23^

In this study, we analysed participants from England, as vaccination data for Scotland and Wales were not available. Although vaccination started in early December 2020 in the UK, we restricted our analyses to periods when both vaccines were rolled out to maximise comparability in individual- and population-level characteristics, including indication for vaccination, ongoing public health restrictions and predominant virus variants. With this in mind, we enrolled one-dose and two-dose Covid-19 vaccine cohorts covering from Jan 11, 2021 to Feb 28, 2021, and from March 22, 2021 to May 9, 2021 respectively. Participants with primary care records generated using the TTP software, which did not contain a specific vaccine type, were excluded.

### Data sources

#### Data linkage

Several external data sources have been linked to UK Biobank to enable Covid-19 research, including primary care electronic health records, hospital admissions data from Hospital Episode Statistics (HES), and Covid-19 tests from Public Health England (PHE).^24^ Data coverage for each data source is listed in the **Supplementary Methods**.

#### Vaccination status

Vaccination status was obtained from GP prescription records, including the date of receipt of each dose and the vaccine brand. We used dm+d codes (Dictionary of medicines and devices used across the UK’s National Health Service) to identify BNT162b2 [*39115611000001103*] and ChAdOx1 [*39114911000001105*] Covid-19 vaccination.

#### Study outcomes

In the UK, people at any age with any of these three Covid-19 symptoms (a high temperature, a new and continuous cough, or loss or change in the sense of smell or taste) are recommended to take a free Polymerase-chain-reaction (PCR) test by ordering a self-swab home PCR test kit or booking an appointment at a walk-in or drive-through test site. People could also access this service if they were at high risk of infection, e.g., close contact with a case (see https://www.gov.uk/get-coronavirus-test for details). All these tests were recorded by PHE and linked to UK Biobank, providing information on test dates and results. In this study, we defined infection as having a positive PCR test for SARS-CoV-2; and Covid-19 hospitalization (infection requiring hospital admission) based on the UKBB derived algorithm described in the **Supplementary Methods**.

#### Covariates

We assessed multiple characteristics potentially associated with Covid-19 risk and/or vaccination, therefore considered as confounders. Study covariates included socio-demographics (age, sex, ethnicity), socio-economic status (index of multiple deprivations, education levels), physical measurements (body mass index), healthcare resource use (prescribed medications and the number of hospital admissions three years before the vaccination date), and comorbidities. The calendar week of receipt of the Covid-19 vaccine was also included as it affected the probability of receiving different vaccines and infection risk (through changes in community transmission level).

### Statistical analyses

The outcome risk assessment window for the one-dose cohort went from receiving the first dose to the earliest of outcome occurrence, receiving the second dose, or 14 weeks after the vaccination. For the two-dose cohort, follow-up was from receiving the second dose to outcome occurrence or end of the study (18/10/2021).

We used the propensity score-based inverse probability of treatment weighting (IPW) to minimise confounding.^25^ The specification of propensity score modelling was described in **Supplementary Methods**. We generated Kaplan-Meier plots to depict the cumulative incidence of outcome over time in each cohort. We applied Cox proportional hazards regression with robust variance estimators to derive average hazard ratio (HR) and calculated incidence rates using weighted counts and follow-up time. We assessed the proportionality of hazards in the Cox models by visually inspecting scaled Schoenfeld residuals.

To evaluate for potential heterogeneity of the comparative effectiveness among specific demographic subgroups and overtime after the second dose, we performed several secondary analyses by including multiplicative interaction terms between the vaccine types and the following categories separately: age (50-75 years or > 75 years), sex (male or female), ethnicity (white or other ethnic groups), BMI (< 25 vs &#x2265; 25), and four weeks’ consecutive time intervals.

We conducted the negative control experiment to assess potential residual (unobserved) confounding. Three clinical outcomes (limb pain, fracture, and peptic ulcer events) were pre-specified and should not be associated with vaccination status (BNT162b2 vs ChAdOx1) if potential confounders have been adequately controlled for.

Finally, we performed a sensitivity analysis using propensity score 1:1 matching without replacement. Specifically, we set a caliper of width equal to 0.2 of the standard deviation of the logit of the propensity score.^26^ We used R version 3.5.1 for all analyses.

## Data Availability

All data produced in the present study are available upon reasonable request to the authors

## Supplementary items

**Supplementary Figure 1:**
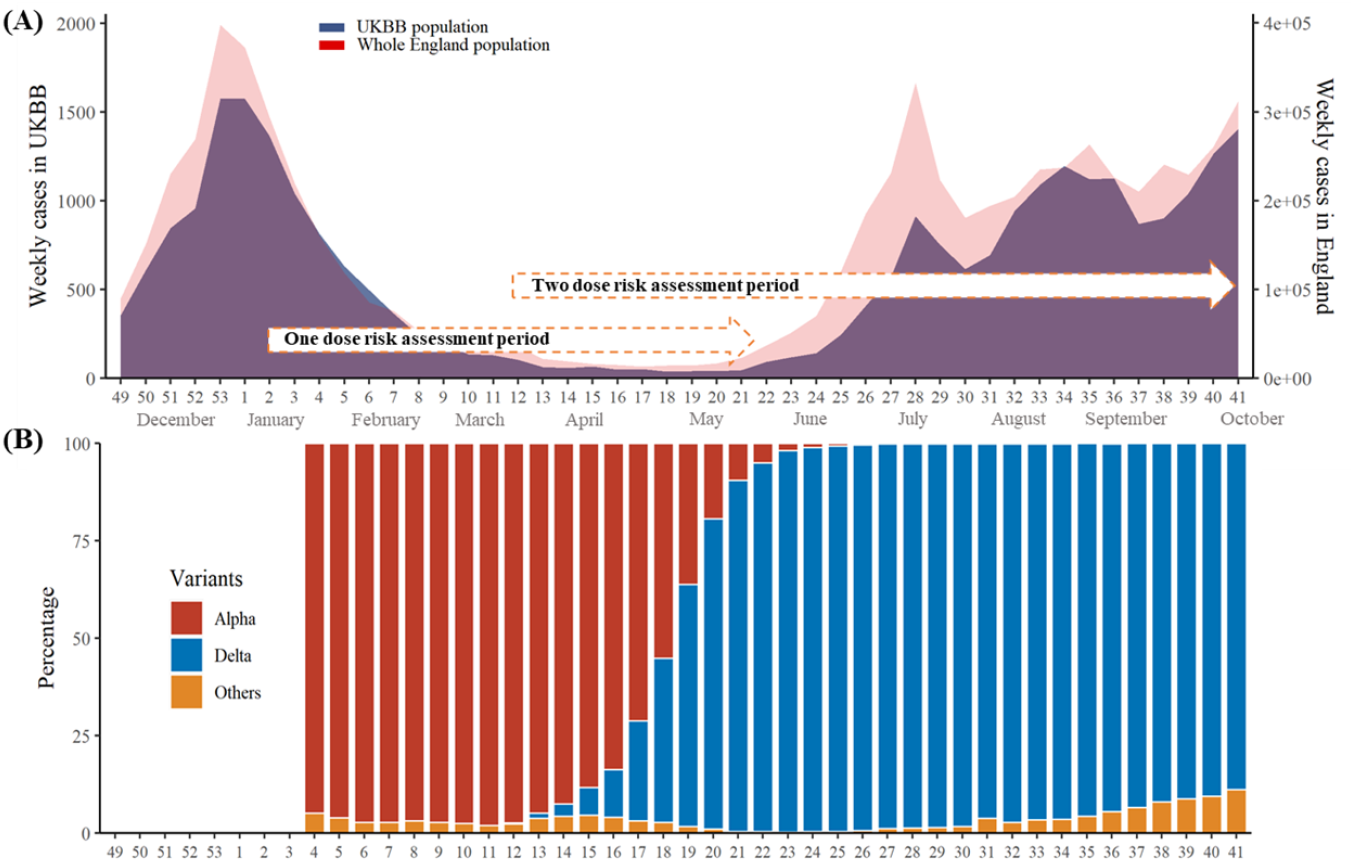
The transmission dynamics of SARS-CoV-2 (A) and its dominant variants (B) during the study period. Notes: The data for weekly Covid-19 cases and SARS-CoV-2 variants of concern in England were from the Public Health England (https://coronavirus.data.gov.uk/).

**Supplementary Table 2:** Baseline characteristics of the one dose cohorts.

|  | Before weighting |  | After weighting |  |
| --- | --- | --- | --- | --- |
|  | BNT162b2 | ChAdOx1 | BNT162b2 | ChAdOx1 |
| Number | 70097 | 98551 | 167938 | 167917 |
| **Previous infection (%)** | 1843 ( 2.6) | 2649 ( 2.7) | 4635.7 ( 2.8) | 4662.1 ( 2.8) |
| **Vaccination calendar week (%)** |  |  |  |  |
| Week2 | 12224 (17.4) | 3127 ( 3.2) | 15716.6 ( 9.4) | 14681.5 ( 8.7) |
| Week3 | 15591 (22.2) | 15728 (16.0) | 31194.2 (18.6) | 31220.5 (18.6) |
| Week4 | 14783 (21.1) | 16990 (17.2) | 32056.9 (19.1) | 32096.3 (19.1) |
| Week5 | 7655 (10.9) | 24264 (24.6) | 31629.9 (18.8) | 31957.0 (19.0) |
| Week6 | 7573 (10.8) | 19492 (19.8) | 26858.2 (16.0) | 27070.1 (16.1) |
| Week7 | 5907 ( 8.4) | 11282 (11.4) | 16731.3 (10.0) | 17059.5 (10.2) |
| Week8 | 6364 ( 9.1) | 7668 ( 7.8) | 13751.8 ( 8.2) | 13832.5 ( 8.2) |
| **Demographics** |  |  |  |  |
| Age, mean (sd) | 71.35 (7.21) | 71.06 (6.02) | 71.09 (6.68) | 71.11 (6.48) |
| Sex, male (%) | 31203 (44.5) | 43432 (44.1) | 74111.1 (44.1) | 74156.6 (44.2) |
| BMI, mean (sd) | 27.56 (4.78) | 27.46 (4.76) | 27.51 (4.77) | 27.51 (4.78) |
| **Ethnicity (%)** |  |  |  |  |
| Asian or Asian British | 1479 ( 2.1) | 1563 ( 1.6) | 3112.1 ( 1.9) | 3085.9 ( 1.8) |
| Black or Black British | 1136 ( 1.6) | 1138 ( 1.2) | 2311.8 ( 1.4) | 2220.0 ( 1.3) |
| Chinese | 224 ( 0.3) | 263 ( 0.3) | 478.5 ( 0.3) | 481.6 ( 0.3) |
| Do not know | 30 ( 0.0) | 41 ( 0.0) | 71.5 ( 0.0) | 75.7 ( 0.0) |
| Mixed | 397 ( 0.6) | 460 ( 0.5) | 851.1 ( 0.5) | 840.7 ( 0.5) |
| Other ethnic group | 657 ( 0.9) | 703 ( 0.7) | 1374.9 ( 0.8) | 1363.6 ( 0.8) |
| Prefer not to answer | 2248 ( 3.2) | 3102 ( 3.1) | 5267.6 ( 3.1) | 5257.8 ( 3.1) |
| White | 63926 (91.2) | 91281 (92.6) | 154471.3 (92.0) | 154592.2 (92.1) |
| **Education (%)** |  |  |  |  |
| None of the above | 12247 (17.5) | 16502 (16.7) | 28750.7 (17.1) | 28763.7 (17.1) |
| Prefer not to answer | 2101 ( 3.0) | 2323 ( 2.4) | 4455.3 ( 2.7) | 4320.0 ( 2.6) |
| College or university degree | 21484 (30.6) | 31157 (31.6) | 52075.8 (31.0) | 52190.1 (31.1) |
| A levels as levels or equivalent | 7498 (10.7) | 11054 (11.2) | 18481.5 (11.0) | 18474.9 (11.0) |
| O levels gcs es or equivalent | 14812 (21.1) | 20890 (21.2) | 35573.0 (21.2) | 35668.3 (21.2) |
| Cs es or equivalent | 3473 ( 5.0) | 4776 ( 4.8) | 8245.5 ( 4.9) | 8268.9 ( 4.9) |
| Nvq or hnd or hnc or equivalent | 4605 ( 6.6) | 6501 ( 6.6) | 11099.1 ( 6.6) | 11120.0 ( 6.6) |
| Other professional qualifications | 3877 ( 5.5) | 5348 ( 5.4) | 9257.9 ( 5.5) | 9111.5 ( 5.4) |
| **Indices of Multiple Deprivation, mean (sd)** |  |  |  |  |
| Townsend deprivation index | 17.70 (13.96) | 16.49 (13.37) | 17.08 (13.57) | 17.01 (13.73) |
| Income score | 0.12 (0.10) | 0.11 (0.09) | 0.11 (0.09) | 0.11 (0.10) |
| Employment score | 0.09 (0.06) | 0.08 (0.06) | 0.09 (0.06) | 0.09 (0.06) |
| Health score | -0.11 (0.89) | -0.18 (0.88) | -0.13 (0.89) | -0.14 (0.89) |
| Education score | 14.61 (14.90) | 14.13 (14.83) | 14.40 (14.74) | 14.39 (15.02) |
| Housing score | 19.94 (9.95) | 19.80 (9.98) | 19.70 (9.84) | 19.69 (9.97) |
| Crime score | -0.04 (0.78) | -0.09 (0.77) | -0.07 (0.77) | -0.07 (0.77) |
| **Medications (past three years)** |  |  |  |  |
| Lipid lowering drugs | 33093 (47.2) | 43266 (43.9) | 76151.4 (45.3) | 75957.6 (45.2) |
| RAS inhibitors | 21956 (31.3) | 29023 (29.4) | 50602.1 (30.1) | 50609.0 (30.1) |
| Other anti-hypertensives | 11158 (15.9) | 14517 (14.7) | 25459.6 (15.2) | 25458.1 (15.2) |
| Proton pump inhibitors | 30579 (43.6) | 41572 (42.2) | 72068.6 (42.9) | 71994.1 (42.9) |
| Diabetes medicines | 6666 ( 9.5) | 8288 ( 8.4) | 14982.9 ( 8.9) | 14899.5 ( 8.9) |
| Antidepressants | 14515 (20.7) | 20199 (20.5) | 34836.6 (20.7) | 34987.6 (20.8) |
| Systemic glucocorticoids | 9282 (13.2) | 12919 (13.1) | 22129.9 (13.2) | 22245.6 (13.2) |
| Antithrombotic | 11799 (16.8) | 15061 (15.3) | 26856.6 (16.0) | 26832.6 (16.0) |
| Anticoagulants | 3110 ( 4.4) | 4020 ( 4.1) | 7099.6 ( 4.2) | 7130.8 ( 4.2) |
| Immunosuppressants excl corticosteroids | 1447 ( 2.1) | 2017 ( 2.0) | 3448.5 ( 2.1) | 3492.0 ( 2.1) |
| Antineoplastic agents | 180 ( 0.3) | 251 ( 0.3) | 413.7 ( 0.2) | 413.4 ( 0.2) |
| **Hospital admission frequency (past three years)** | 1.38 (5.52) | 1.34 (5.25) | 1.37 (6.36) | 1.37 (5.12) |
| **Charlson comorbidity components (past years)** |  |  |  |  |
| Diabetes (uncomplicated) | 9193 (13.1) | 11657 (11.8) | 20797.9 (12.4) | 20800.9 (12.4) |
| COPD | 14156 (20.2) | 19433 (19.7) | 33547.1 (20.0) | 33504.9 (20.0) |
| Chronic kidney disease | 7449 (10.6) | 9368 ( 9.5) | 16714.7 (10.0) | 16713.1 (10.0) |
| Congestive heart failure | 1473 ( 2.1) | 1893 ( 1.9) | 3344.1 ( 2.0) | 3370.8 ( 2.0) |
| Cancer | 10807 (15.4) | 14483 (14.7) | 25028.3 (14.9) | 24975.7 (14.9) |
| Dementia | 681 ( 1.0) | 959 ( 1.0) | 1731.3 ( 1.0) | 1827.9 ( 1.1) |
| Cerebrovascular disease | 2799 ( 4.0) | 3498 ( 3.5) | 6356.5 ( 3.8) | 6324.4 ( 3.8) |
| Diabetes (end-organ damage) | 3821 ( 5.5) | 4755 ( 4.8) | 8556.7 ( 5.1) | 8526.5 ( 5.1) |
| Peripheral vascular disease | 1459 ( 2.1) | 1857 ( 1.9) | 3276.3 ( 2.0) | 3287.2 ( 2.0) |
| Liver disease (moderate to severe) | 279 ( 0.4) | 384 ( 0.4) | 631.3 ( 0.4) | 646.1 ( 0.4) |
| Peptic ulcer | 2151 ( 3.1) | 2877 ( 2.9) | 5081.0 ( 3.0) | 5018.4 ( 3.0) |
| Rheumatoid arthritis | 3118 ( 4.4) | 4177 ( 4.2) | 7266.1 ( 4.3) | 7265.0 ( 4.3) |
| Myocardial infarction | 2573 ( 3.7) | 3254 ( 3.3) | 5890.7 ( 3.5) | 5866.9 ( 3.5) |
| AIDS | 137 ( 0.2) | 183 ( 0.2) | 320.8 ( 0.2) | 310.5 ( 0.2) |
| Malignant cancer | 370 ( 0.5) | 502 ( 0.5) | 859.2 ( 0.5) | 836.4 ( 0.5) |
| Liver disease (mild) | 452 ( 0.6) | 650 ( 0.7) | 1105.4 ( 0.7) | 1110.2 ( 0.7) |
| Hemiplegia | 98 ( 0.1) | 147 ( 0.1) | 263.2 ( 0.2) | 250.8 ( 0.1) |

**Supplementary Table 3:** Baseline characteristics of the two dose cohorts.

|  | Before weighting |  | After weighting |  |
| --- | --- | --- | --- | --- |
|  | BNT162b2 | ChAdOx1 | BNT162b2 | ChAdOx1 |
| Number | 67813 | 89030 | 156301 | 156952 |
| **Previous infection (%)** | 2539 ( 2.9) | 1818 ( 2.7) | 4518.5 ( 2.9) | 4498.7 ( 2.9) |
| **Vaccination calendar week (%)** |  |  |  |  |
| Week12 | 8299 (12.2) | 3972 ( 4.5) | 12366.1 ( 7.9) | 12606.7 ( 8.0) |
| Week13 | 13558 (20.0) | 4373 ( 4.9) | 18073.8 (11.6) | 17812.4 (11.3) |
| Week14 | 12976 (19.1) | 13900 (15.6) | 26803.0 (17.1) | 26791.8 (17.1) |
| Week15 | 13365 (19.7) | 16445 (18.5) | 29957.9 (19.2) | 29935.1 (19.1) |
| Week16 | 6892 (10.2) | 21591 (24.3) | 28278.7 (18.1) | 28468.5 (18.1) |
| Week17 | 7037 (10.4) | 16945 (19.0) | 23698.1 (15.2) | 23941.3 (15.3) |
| Week18 | 5686 ( 8.4) | 11804 (13.3) | 17123.5 (11.0) | 17396.8 (11.1) |
| **Demographics** |  |  |  |  |
| Age, mean (sd) | 72.47 (6.97) | 71.51 (5.98) | 71.80 (6.74) | 71.81 (6.18) |
| Sex, male (%) | 29825 (44.0) | 39250 (44.1) | 68880.5 (44.1) | 69199.6 (44.1) |
| BMI, mean (sd) | 27.47 (4.67) | 27.46 (4.74) | 27.50 (4.72) | 27.50 (4.75) |
| **Ethnicity (%)** |  |  |  |  |
| Asian or Asian British | 1279 ( 1.9) | 1348 ( 1.5) | 2656.8 ( 1.7) | 2641.7 ( 1.7) |
| Black or Black British | 924 ( 1.4) | 969 ( 1.1) | 1957.9 ( 1.3) | 1921.4 ( 1.2) |
| Chinese | 185 ( 0.3) | 225 ( 0.3) | 405.8 ( 0.3) | 409.4 ( 0.3) |
| Do not know | 26 ( 0.0) | 37 ( 0.0) | 68.3 ( 0.0) | 66.4 ( 0.0) |
| Mixed | 345 ( 0.5) | 408 ( 0.5) | 730.0 ( 0.5) | 735.3 ( 0.5) |
| Other ethnic group | 560 ( 0.8) | 613 ( 0.7) | 1182.2 ( 0.8) | 1171.0 ( 0.7) |
| Prefer not to answer | 2143 ( 3.2) | 2737 ( 3.1) | 4807.1 ( 3.1) | 4881.0 ( 3.1) |
| White | 62351 (91.9) | 82693 (92.9) | 144492.9 (92.4) | 145126.1 (92.5) |
| **Education (%)** |  |  |  |  |
| None of the above | 12648 (18.7) | 15435 (17.3) | 28036.2 (17.9) | 28234.6 (18.0) |
| Prefer not to answer | 2031 ( 3.0) | 2116 ( 2.4) | 4097.4 ( 2.6) | 4059.5 ( 2.6) |
| College or university degree | 20513 (30.2) | 27567 (31.0) | 47638.2 (30.5) | 47836.8 (30.5) |
| A levels as levels or equivalent | 7069 (10.4) | 9848 (11.1) | 16888.3 (10.8) | 16951.9 (10.8) |
| O levels gcs es or equivalent | 14183 (20.9) | 19030 (21.4) | 33181.6 (21.2) | 33314.8 (21.2) |
| Cs es or equivalent | 2970 ( 4.4) | 4142 ( 4.7) | 7166.0 ( 4.6) | 7180.5 ( 4.6) |
| Nvq or hnd or hnc or equivalent | 4440 ( 6.5) | 5916 ( 6.6) | 10415.2 ( 6.7) | 10483.2 ( 6.7) |
| Other professional qualifications | 3959 ( 5.8) | 4976 ( 5.6) | 8878.2 ( 5.7) | 8891.1 ( 5.7) |
| Indices of Multiple Deprivation, mean (sd) |  |  |  |  |
| Townsend deprivation index | 17.35 (13.69) | 16.39 (13.27) | 17.00 (13.51) | 16.97 (13.62) |
| Income score | 0.11 (0.09) | 0.11 (0.09) | 0.11 (0.09) | 0.11 (0.09) |
| Employment score | 0.09 (0.06) | 0.08 (0.06) | 0.09 (0.06) | 0.09 (0.06) |
| Health score | -0.12 (0.89) | -0.18 (0.88) | -0.14 (0.88) | -0.14 (0.88) |
| Education score | 14.39 (14.77) | 14.10 (14.80) | 14.35 (14.72) | 14.35 (14.96) |
| Housing score | 19.85 (9.90) | 19.66 (9.92) | 19.64 (9.80) | 19.66 (9.97) |
| Crime score | -0.05 (0.78) | -0.09 (0.77) | -0.07 (0.77) | -0.07 (0.77) |
| **Medications (past three years)** |  |  |  |  |
| Lipid lowering drugs | 33138 (48.9) | 40248 (45.2) | 73220.9 (46.8) | 73457.8 (46.8) |
| RAS inhibitors | 21768 (32.1) | 26794 (30.1) | 48369.8 (30.9) | 48521.5 (30.9) |
| Other anti-hypertensives | 11297 (16.7) | 13489 (15.2) | 24636.9 (15.8) | 24732.6 (15.8) |
| Proton pump inhibitors | 30074 (44.3) | 38128 (42.8) | 68201.9 (43.6) | 68396.9 (43.6) |
| Diabetes medicines | 6168 ( 9.1) | 7556 ( 8.5) | 13866.2 ( 8.9) | 13854.3 ( 8.8) |
| Antidepressants | 13877 (20.5) | 18277 (20.5) | 32391.9 (20.7) | 32511.1 (20.7) |
| Systemic glucocorticoids | 9126 (13.5) | 11585 (13.0) | 20797.3 (13.3) | 20784.2 (13.2) |
| Antithrombotic | 11903 (17.6) | 14077 (15.8) | 26024.1 (16.6) | 26049.1 (16.6) |
| Anticoagulants | 3392 ( 5.0) | 3969 ( 4.5) | 7289.0 ( 4.7) | 7271.8 ( 4.6) |
| Immunosuppressants excl corticosteroids | 1357 ( 2.0) | 1823 ( 2.0) | 3216.2 ( 2.1) | 3222.4 ( 2.1) |
| Antineoplastic agents | 185 ( 0.3) | 232 ( 0.3) | 411.4 ( 0.3) | 398.8 ( 0.3) |
| **Hospital admission frequency (past three years)** | 1.39 (5.82) | 1.32 (5.02) | 1.37 (6.36) | 1.36 (4.89) |
| **Charlson comorbidity components (past years)** |  |  |  |  |
| Diabetes (uncomplicated) | 8663 (12.8) | 10667 (12.0) | 19443.3 (12.4) | 19495.5 (12.4) |
| COPD | 13737 (20.3) | 17715 (19.9) | 31566.4 (20.2) | 31582.3 (20.1) |
| Chronic kidney disease | 7746 (11.4) | 8924 (10.0) | 16513.0 (10.6) | 16558.8 (10.6) |
| Congestive heart failure | 1548 ( 2.3) | 1814 ( 2.0) | 3355.0 ( 2.1) | 3359.2 ( 2.1) |
| Cancer | 10937 (16.1) | 13601 (15.3) | 24305.2 (15.6) | 24366.2 (15.5) |
| Dementia | 820 ( 1.2) | 1004 ( 1.1) | 1871.1 ( 1.2) | 1942.2 ( 1.2) |
| Cerebrovascular disease | 2860 ( 4.2) | 3327 ( 3.7) | 6199.3 ( 4.0) | 6173.1 ( 3.9) |
| Diabetes (end-organ damage) | 3638 ( 5.4) | 4411 ( 5.0) | 8099.5 ( 5.2) | 8084.2 ( 5.2) |
| Peripheral vascular disease | 1513 ( 2.2) | 1760 ( 2.0) | 3258.4 ( 2.1) | 3256.7 ( 2.1) |
| Liver disease (moderate to severe) | 258 ( 0.4) | 348 ( 0.4) | 605.6 ( 0.4) | 606.2 ( 0.4) |
| Peptic ulcer | 2190 ( 3.2) | 2684 ( 3.0) | 4912.0 ( 3.1) | 4898.0 ( 3.1) |
| Rheumatoid arthritis | 3225 ( 4.8) | 3939 ( 4.4) | 7172.6 ( 4.6) | 7153.7 ( 4.6) |
| Myocardial infarction | 2541 ( 3.7) | 3053 ( 3.4) | 5663.6 ( 3.6) | 5663.5 ( 3.6) |
| AIDS | 131 ( 0.2) | 164 ( 0.2) | 294.3 ( 0.2) | 288.0 ( 0.2) |
| Malignant cancer | 352 ( 0.5) | 462 ( 0.5) | 789.2 ( 0.5) | 804.3 ( 0.5) |
| Liver disease (mild) | 421 ( 0.6) | 579 ( 0.7) | 1005.7 ( 0.6) | 1022.9 ( 0.7) |
| Hemiplegia | 90 ( 0.1) | 123 ( 0.1) | 225.5 ( 0.1) | 222.0 ( 0.1) |

**Supplementary Table 4:** Sensitivity analysis (propensity score matching)

|  | BNT162b2 vs ChAdOx1 |  |  |  |
| --- | --- | --- | --- | --- |
|  | Number | Events | Incidence | HRs |
| After first-dose |  |  |  |  |
| Covid-19 infection | 58676/ 58676 | 154/ 193 | 12.6/ 15.7 | 0.80 (0.65 - 0.99) |
| Covid-19 hospitalization | 58676/ 58676 | 29/ 32 | 2.36/ 2.60 | 0.91 (0.55 - 1.51) |
| After second-dose |  |  |  |  |
| Covid-19 infection | 54422/ 54422 | 1089/ 1582 | 39.5/ 57.6 | 0.69 (0.63 - 0.74) |
| Covid-19 hospitalization | 54422/ 54422 | 93/ 142 | 3.35/ 5.13 | 0.65 (0.50 - 0.85) |

## Supplementary Methods

### Sources and timelines for data availability

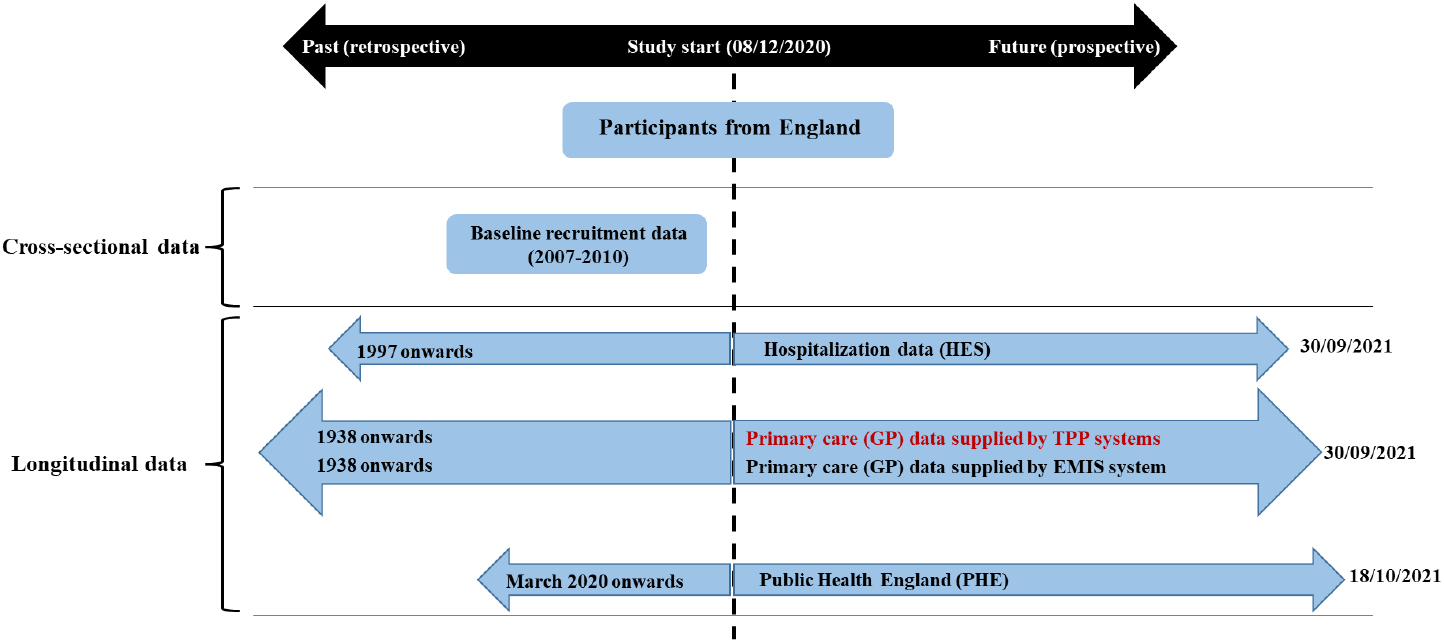

The baseline recruitment data includes detailed demographic, socioeconomic, lifestyle factors, and physical activities information. The HES data includes hospital administration, operations and diagnoses information. The GP data includes diagnoses and prescription information. Of note, the Covid-19 vaccination has been integrated into EMIS GP’s prescription records but not into the TPP. The PHE data includes Covid-19 test results.

### Construction of the inpatient indicator for UKBB participants

The construction of the “origin” field is based on information provided on the specimen request form. If the specimen was marked as being from an acute (emergency) care provider, an A&E department, an inpatient location, or resulted from health care associated infection, it is recorded by PHE as an inpatient sample. Tests marked as being from “Healthcare Worker Testing” are never recorded as inpatient samples, though some may also carry an acute flag.

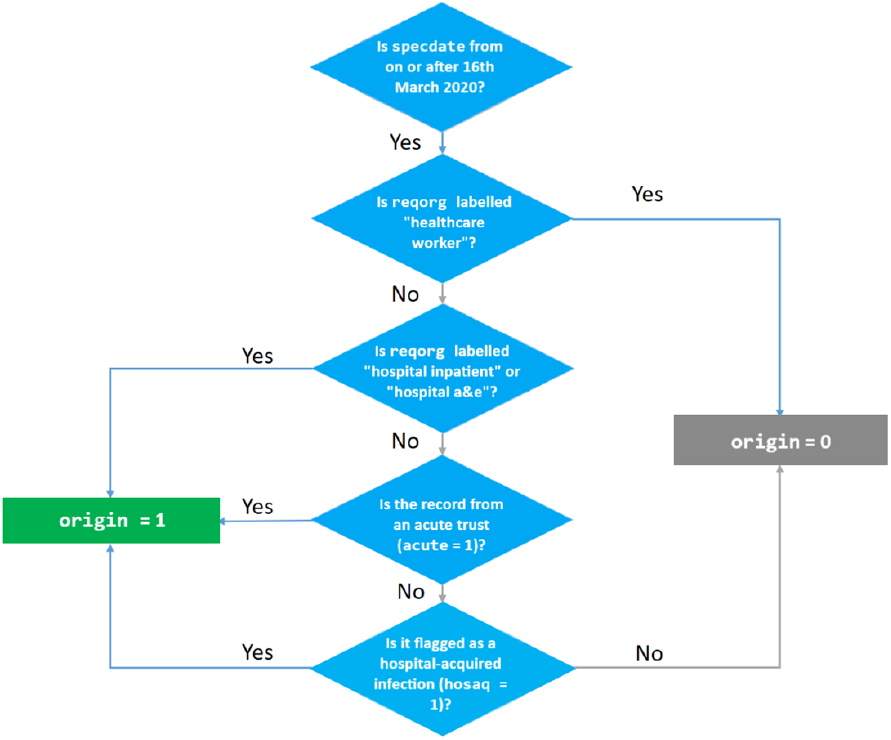

The aim of designating inpatient status for the SARS-CoV-2 test was to indicate severity of Covid-19 infection. SARS-CoV-2 tests taken in hospital can be undertaken for several reasons including symptomatic patients requiring hospital admission, or general inpatient screening, which includes asymptomatic patients. Furthermore, the algorithm used to flag inpatient status may not necessarily indicate inpatient care in all cases. For example, some tests flagged as coming from “acute” trusts will likely not be inpatients, since these trusts may also perform tests on behalf of GPs and others, and tests requested by A&E may be for patients who are not then admitted.

The flow chart below illustrates the logic used by PHE to generate an indicator of whether a test result was obtained from a hospital inpatient or not (depicted as the “origin” field in the covid19_result table). The fields used to construct the “origin” field have also been released to enable researchers to replicate it and construct their own alternatives if desired.

PHE’s designation of inpatient status can be compared to hospital episodes statistics (HES) dates of admission and discharge made by NHS trusts. A comparison between SARS-CoV-2 positive inpatient status versus inpatient diagnosis codes (ICD-10 diagnosis codes U071 or U072) for Covid-19 (from HES) can also be made. For further details, see the following webpage: https://news.bugbank.uk/2020/08/identifying-inpatients-comparison-to.html.

### Propensity score modelling and inverse probability of treatment weighting

We calculated propensity scores for a vaccination with BNT162b2 against ChAdOx1 using logistic regression. The variables included in the model were previous Covid-19 infection status (binary), age on the vaccination date (continuous linear), sex (binary), ethnicity (categorial, collected at the UK Biobank recruitment), multiple socio-economic deprivation scores (Townsend deprivation index, income score, employment score, health score, education score, housing score, and crime score; continuous linear, collected at the UK Biobank recruitment), education levels (categorial, collected at the UK Biobank recruitment), body mass index (continuous linear, collected at the UK Biobank recruitment), a list of pre-specified medications (binary, obtained from primary care prescription records), the number of hospital admissions (continuous linear, obtained from HES), and comorbidities (binary, obtained from primary care diagnosis records). The weights for each participant were then computed based on the Rosenbaum formula: 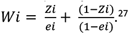 ^27^ To reduce unstable estimate due to extreme weights in the tails of the propensity score distribution, we used asymmetric trimming to exclude people whose propensity score was below the 1^th^ percentile of the propensity score of the BNT162b2cohort and above the 99^th^ percentile of the propensity score of the ChAdOx1 cohort.^28^

## Acknowledgements and funding

DPA is funded through an NIHR Senior Research Fellowship (Grant number SRF-2018-11-ST2-004). The views expressed in this publication are those of the author(s) and not necessarily those of the NHS, the National Institute for Health Research or the Department of Health. Mr Junqing Xie has been awarded a Jardine-Oxford Graduate Scholarship and a titular Clarendon Fund Scholarship outside the submitted work. AP-U is supported by the Medical Research Council (grant numbers MR/K501256/1, MR/N013468/1).

